# The expression alteration of *Golim4* (Gpp130) and its prognostic value in epithelial ovarian cancer

**DOI:** 10.1101/2024.10.01.24314617

**Authors:** Sadaf Valeh Sheida, Akram Gholipour, Hossein Bayat Varchaghi, Seyed Javad Mowla

## Abstract

**Background:** Among common cancers in women, ovarian cancer has the lowest incidence; however, mortality rates are more than other gynecological cancers due to the lack of specific symptoms during early stages and absence of diagnostic markers. Golgi integral membrane protein 4 (*GOLIM4*) is a member of transporter complex which is located to the cis/medial of Golgi, and its role in tumor is unknown. In this study we aimed to assess the expression of *Golim4* (GPP130) in epithelial ovarian cancer (EOC) patients.

**Methods:** The expression levels of *Golim4* in 58 malignant EOC specimens and 11 benign and normal control tissues were determined by real-time RT-PCR. Clinicopathological characteristics were analyzed. Non-parametric survival analysis was estimated by using Kaplan-Meier and log-rank test. Parametric survival analysis was evaluated through multivariate Cox regression analysis and generalized Gamma survival model.

**Result:** Our findings showed that *Golim4* was significantly down-regulated in Serous Papillary type of EOC tissue specimens than in non-cancerous tissues (P<0.001); however; the expression of *Golim4* in total EOC tissue specimens revealed non-significant up-regulation. Kaplan-Meier analysis and log-rank test have suggested that EOC patients with low *Golim4* expression have shorter overall survival when compared with patients with other expression groups (log-rank test P<0.05). Semi-parametric Cox proportional hazards model indicated that the status of *Golim4* were independent predictor of overall survival in patients with EOC. Correlation analysis has been done by Spearman Rank correlation coefficient.

## Introduction

Epithelial ovarian cancer (EOC) is the fifth leading cause of cancer deaths and the most lethal gynecological malignancy. Globally, it was estimated that 295.414 women were diagnosed with ovarian cancer and 184,799 women died from this disease in 2018 [1]. The early stages of ovarian cancer have no specific symptoms and there are currently no accurate diagnostic tests for early diagnosis of ovarian cancer. 70% of patients are diagnosed at the late stages where the prognosis of EOC is almost impossible [2]. The huge number of scientific studies in recent years established a new understanding of the biology and structure of tumors in ovarian cancer. New classification of EOC tumors introduced from 2014 by the Federation of Gynecology and Obstetrics (FIGO) and WHO which was based on histopathological findings and the recognition of the exact type of tumor. This categorization has a positive effect on patient prognosis and treatment. According to new category the tumors of the ovary are divided into five groups: serous, Mucinous, Endometroid, Clear cell and Brenner [3]. Serous Papillary Carcinoma is one of the most common epithelial tumors which include 40-50% of epithelial ovarian carcinomas [4]. Copy number variation (CNV) is one of the most common genetic changes in human tumors. Studies using Comparative genomic hybridization (CGH) have revealed that CNV accrued in 36-51% of 3q26 region in epithelial ovarian patients and Amplification of the 3q26 locus is one of the most common alterations, observed in 85% of epithelial ovarian cancer cases [5]. According to these observations, we analyzed the cBioPortal for Cancer Genomics Database. According to the studies of TCGA in cBio Cancer Genomics Portal (http://www.cbioportal.org), on 585 individuals with epithelial ovarian cancer, one region at 3q26 found to be increased in copy number in approximately 20% of patients, contains *Golim4*[6]. *Golim4 (Golgi Integral Membrane Protein 4)* is a protein coding gene, encodes phosphorylated and glycosylated protein which located inside the Cis/Medial Golgi membrane with a weight of 130 kDa. The sequence of the cDNA encoding this protein has shown that this protein is a type 2 membrane protein and involved in processing and trafficking between the *cis* Golgi and endosomes [7]. The position of the *Golim4* gene is located on chromosome 3 (3q26.2). As a result, the *Golim4* gene is one of the genes in this controversial region that has led us to detect changes in the expression of this gene in epithelial ovarian cancer. The present study was conducted to evaluate the clinical significance of *Golim4*.

## Materials and methods

### Samples collection

Sixty-eight tissue samples were obtained from surgical specimens at Hajar and Imam Khomeini Hospitals (Tehran, Iran) between 2008 and 2016. We used two groups in this study. The first group were patients with cancerous changes. Histology cancer patient groups were as follows: Endometroid Adenocarcinoma (n=8), Serous Papillary Carcinoma (n=23), Serous Cystadenocarcinoma (n=1), Serous Adenocarcinoma (n=1), Mucinous Adenocarcinoma (n=1), Mucinous Cystadenoma (n=3), Clear Cell Adenocarcinoma (n=7), Small Cell Carcinoma (n=2), LMP2 (n=7), Undifferentiated Carcinoma (n=1), Poorly Differentiated Carcinoma (n=2), not otherwise specified (NOS) (n=1). The second group were patients with non-cancerous changes (n=11). The patient’s ages were between 28 to 77 years with a mean age of 53 years. Tissues were snap-frozen in liquid nitrogen after excision, transported to the Pathology Laboratory, and stored at -80°C. The diagnosis and the histological grading were approved by pathologists. Written informed consent was collected from all patients. The median follow-up time was obtained to be 40 months (range 7-90 months). We defined overall survival (OS) based on the elapsed time from the end of surgery to the death.

### RNA extraction

Tissue specimens were snap-frozen in liquid nitrogen and stored at−80°C. Moreover, the patients’ Total RNA was extracted from frozen samples using TRIzol reagent (Ambion, Carlsbad, CA), according to the manufacturer’s instructions. To avoid any genomic DNA contamination, RNAs were treated with DNaseI (Fermentas, Lithuania). For evaluating the quality of RNA, we used gel electrophoresis and the concentration of RNA was measured by spectrophotometer at 260 nm.

### CDNA synthesis and RT-PCR

Complementary DNA (cDNA) was synthesized from 1 μg total RNA for tissue samples, using random hexamer and oligo dT primers via Prime Script RT (Takara, Japan), according to the manufacturer’s instructions. PCR was performed using 1 μg of synthesized cDNA with 2 μL of 10× PCR buffer (Ampliqon, Denmark), 0.5 μL of each primer and deionized distilled water in a 10 μL PCR reaction volume. The PCR was carried out for 40 cycles for all genes, except for the internal control gene, Act B, where the number of cycles was 25.

### Quantitative real-time PCR

Quantitative real-time PCR was performed by Applied Biosystems StepOne Real-Time PCR System (ABI). A total 15 μL of reaction mixture involved 1 μL of cDNA template, 0.5 μL forward and reverse primers, and 7.5 μL of Biofact 2X SYBR Green PCR Mix (Takara, Japan) was amplified according to following instruction: initial denaturation at 95°C for 5 min, followed by 40 cycles of denaturation at 95°C for 10 second, and annealing at 62°C for 20 second and extension at 72°C for 18 second. Each sample was duplicated in Quantitative Real-Time PCR. GAPDH used as an internal control and the level of GAPDH expression was measured in all samples to normalize mRNA levels for the differences in sample concentration and loading. The sequences of the primers were forward primer: ‘5ATGGGGAAGGTGAAGGTCG3’ and reverse primer: ‘5GGGGTCATTGATGGCAACAATA3’ for GAPDH and forward primer: ‘5TTAGGTGGCAACAAAGTCGTG 3’ and reverse primer: ‘5TCCTTCTTCCCACTTTTTGGC 3’ for *Golim4*. The relative expression of each gene was performed by the comparative threshold cycle as described by GraphPad Prism 6 Demo and data analysis was conducted using the comparative CT method. Relative expression of each gene was calculated by 2^−ΔΔCt^ formula.

### Statistical Methods

According to the results of parametric assumptions, the two-independent samples t-test or Mann-Whitney non-parametric test was performed to compare the central tendency (e.g., mean for normal and median for non-normal variables) between the groups by GraphPad Prism version 6. Furthermore, associations between *Golim4* expression and the clinical pathological characteristics were evaluated by the chi-square test and Spearman rank correlation analysis. Survival analysis was done by non-parametric analysis (e.g., log-rank test), semi-parametric analysis (e.g., cox PH regression), and parametric analysis (e.g., Generalized Gamma model). Survival curves were plotted using Kaplan-Meier method. All of survival analysis was done using STATA version 14.2 (College Station, Texas 77845 USA). P-value less than 0.05 were considered as statistically significant.

## Result

RT-PCR used to set up the optimal conditions for amplification of the candidate genes in EOC and non-cancerous tissues. RT-PCR analysis showed that *Golim4* has expression in both EOC and non-cancerous tissues (Figure 1A). Expression level of candidate gene was determined by quantitative Real-Time PCR analysis in 57 EOC and 11 non-cancerous specimens. The relative expression of *Golim4* in tumor and non-cancerous specimens failed to show statistically significant differences (Figure 1B). The expression levels of *Golim4* in Serous carcinoma tissues (n= 25) were determined by Real-Time PCR. Our findings showed that *Golim4* was significantly downregulated in serous carcinoma tissue specimens than in non-cancerous tissues (mean ± SD: 5.15 ± 0.88 vs. 12.7± 2.82; P<0.001; Figure1C); however, the relative gene expression of *Golim4* between different stages of tumor samples revealed no significant differences between low and high stage tumors. The patients were categorized into low and high expression groups. Decreased expression of *Golim4* was significantly associated with tumor size (p<0.05). No significant correlation was determined between *Golim4* expression and other clinicopathological factors such as stage and grade.

**Figure 1.**
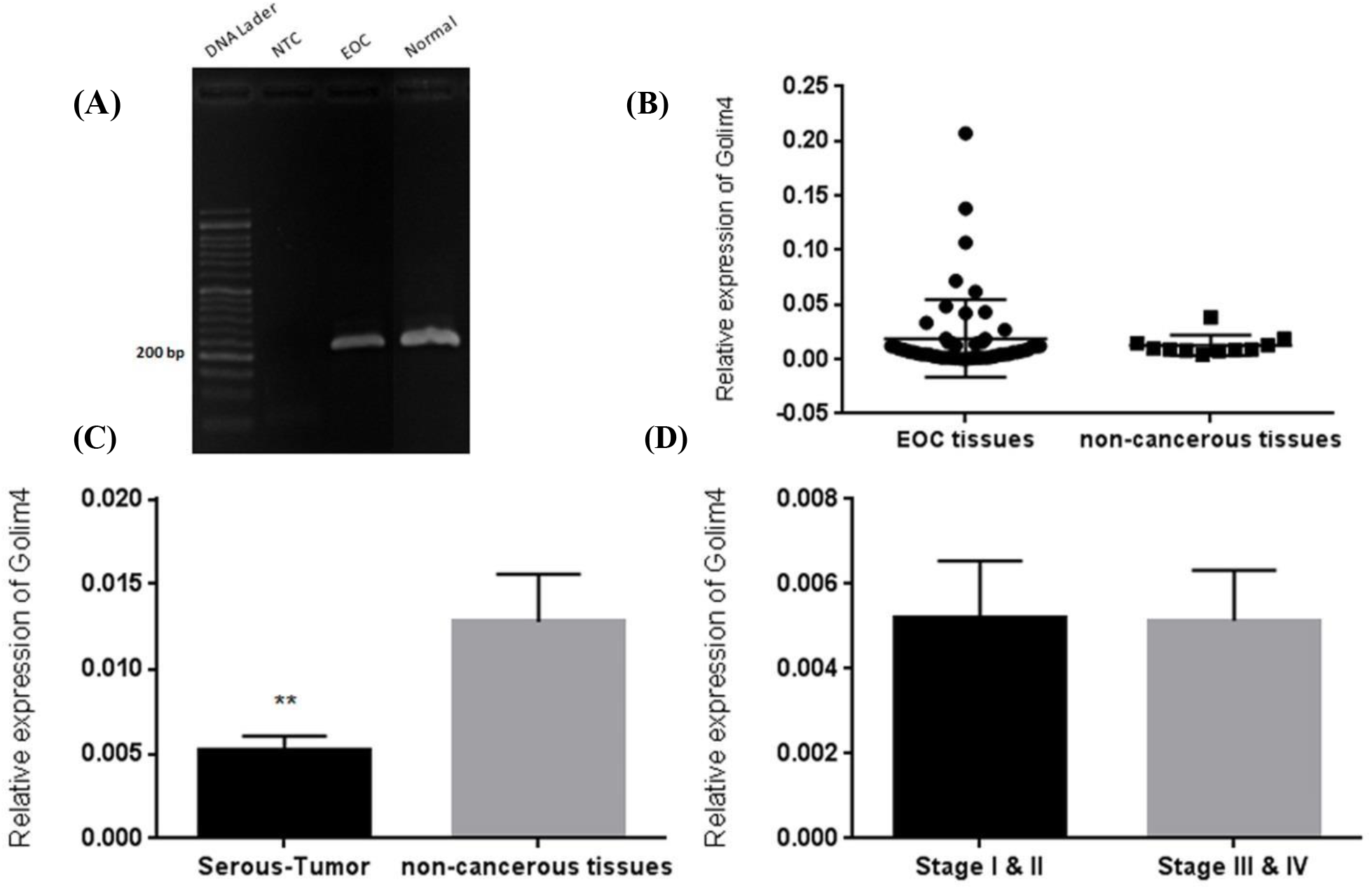
**(A)**: RT-PCR analysis of *Golim4* expression in EOC and non-cancerous tissues. **(B)**: Relative gene expression of *Golim4* in EOC and normal samples. The expression of the gene was normalized against the housekeeping gene *GAPDH* and calculated by Livak method (2^-ΔΔCt^). The result revealed no significant differences between *Golim4* transcripts in EOC and non-cancerous tissues. **(C)**: Expression of *Golim4* in serous tissue specimens and non-cancerous tissues was detected by Real-Time PCR. The result showed that *Golim4* transcripts were significantly downregulated in serous type of ovarian cancer (P = 0.0019). (**D)**: The relative gene expression of *Golim4* between different stages of tumor samples revealed no significant differences between low and high stage tumors.

### The relationship of Golim4 expression with prognosis

Kaplan-Meier analysis and log-rank test have suggested that EOC patients with low *Golim4* expression have shorter overall survival when compared with patients with other expression groups (Figure 2). Univariate and multivariate analyses were used to assess whether the *Golim4* expression levels and clinicopathological parameters were independent prognostic factors of EOC patient outcomes. Multivariate Cox proportional hazards model showed that the low expression of *Golim4* and advanced FIGO stage were independent predictor of overall survival (Figure 2).

**Figure 2:**
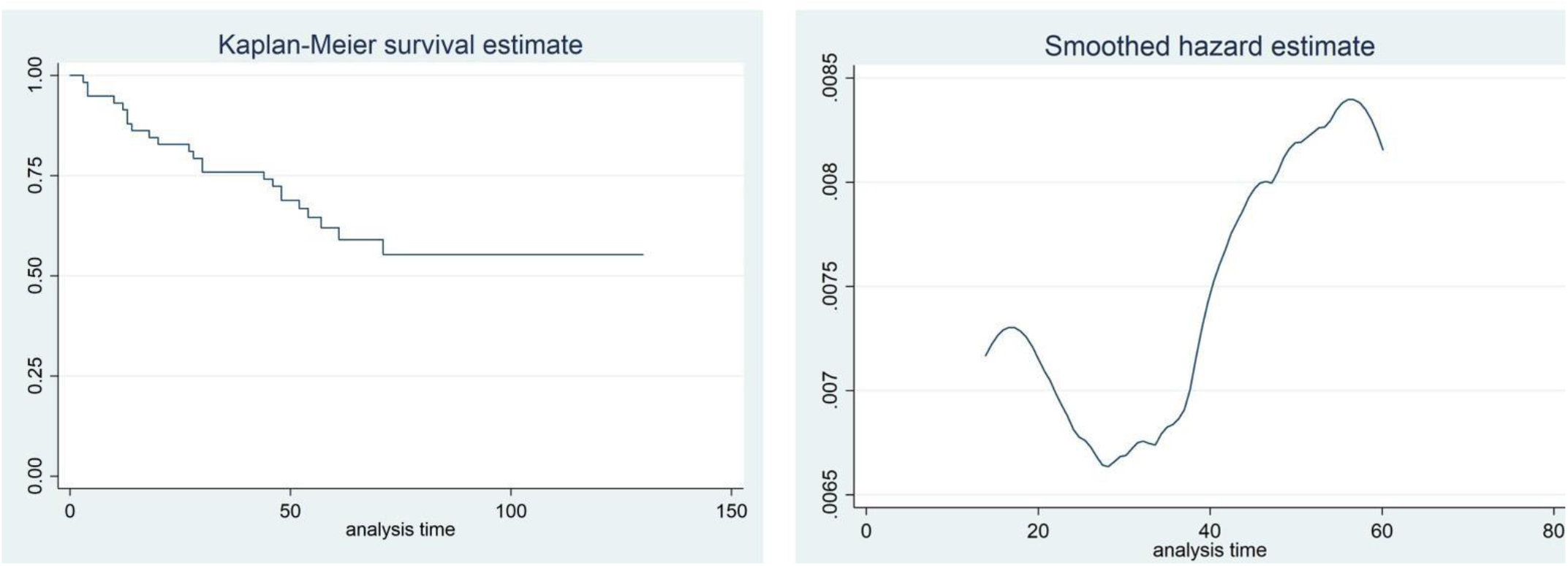
Kaplan-Meier curves for survival time in patients with epithelial ovarian cancer divided according to *Golim4* expression level.

## Discussion

Ovarian cancer has the high rate of mortality according to the lack of signs and symptoms such as pain and bleeding at the early stages of the disease and no specific and sensitive diagnostic markers and methods [8]. Clinical and pathological parameters such as grading and staging tumors provide important information about the cancer, but they are not able to predict the correct behavior of the tumors and their malignant potential. Therefore, accurate and timely diagnosis of the exact biologic behavior of the tumor and the determination of the appropriate treatment strategy is necessary [9]. The CA125 test and transvaginal ultrasound are the only standard and reliable methods for diagnosis of the ovarian tumors. The CA125 marker is used as the most important screening test for ovarian epithelial cancers (EOC). Although CA125 is the only biomarker accepted for ovarian cancer, as a screening test has showed the high percentage of errors and false-positive responses. For example, an increase in the level of this tumor marker is seen in some benign tumors, such as endometriosis and some other cancers such as pancreatic, breast, colon and lung. On the other hand, the biomarker remains unchanged in the 50% of those who are at early stages of the disease [10 - 13]. So, there is still an urgent necessity for finding new biomarkers that could help to detect potentially life-threatening ovarian cancer cases, at a proper and early time. Our candidate gene, Golim4 codes a phosphorylated and glycosylated protein with 130kD. This is a membrane protein that is integrated into the cis/medial Golgi apparatus, which is a kind II Golgi protein. *GOLIM4* is a Golgi membrane protein that circulates between Golgi apparatus and endosomes, and it is pH-sensitive. In fact, the protein produced by this gene has a pH-sensitive lumen which leads to transmission from Golgi to the endosome by altering PH [7]. The probable role of *GOLIM4* in the cell death process was also reported by a number of studies. Based on these studies Mn can downregulate the expression of *GOLIM4* and this process leads to cell death [14, 15]. A change in the number of copies of genes (CNVs) is one of the most common genetic abnormalities in human tumors. Copy number variation (CNVs) can provide the growth benefits to cancer cells through various mechanisms and the creation of invasive phenotypes. Recognition of altered chromosomal regions in solid tumors by Comparative genotypic hybridization (CGH) indicated 90% of chromosomal abnormalities in more than 15 different types of human tumors. In addition, comparative genomic hybridization studies (CGH) on ovarian cancer indicate a wide range of chromosomal changes in the area of 3q26, and the amplification of this region has been observed in 80% of the samples that are present in the early stages of ovarian cancer [5, 16, 17]. According to existence of *Golim4* in this controversial region and the role of *Golim4* in cell death, we have chosen our candidate gene and looked for whether the expression of this gene is changed in tumor specimens in comparison with non-cancerous tissues. Until now, there have been lots of different studies over the expression of *Golim4*; however, few studies have been done on the role of this gene in various cancers. For instance, Lu et al. with General examination of genomic changes in ovarian tumors and glioblastoma with reference to TCGA data, introduces *Golim4* as an exacerbating factor in these cancers [18]. A recent publication about Golim4 and its role in the process of head and neck cancer indicated that Golim4 inhibited the proliferation of head and neck cancer, promote apoptosis and regulate cell cycle progression [19]. Another report by Lin et.al. indicate that *Golim4* expression is under the control of *miR-105-3p* and *Golim4* silencing promote cells proliferation and migration and inhibit the apoptosis in breast cancer cell lines. According to this study *miR-105-3p* could downregulate the expression of *Golim4* at both transcriptional and translational levels [20].

## Conclusion

In this study, we found that the expression level of *Golim4* was significantly down-regulated in serous type of epithelial ovarian cancer tissues, compared to the non-cancerous samples. In contrast, the expression level of *Golim4* was showed no significant changes among all types of EOC in comparison with normal tissues. We assume that *Golim4* is a tumor suppressor gene and *miR-105-3p* which has an oncogenic role, silences *Golim4* expression in cancerous tissue. Kaplan-Meier analysis and log-rank test suggested that EOC patients with down-regulated *Golim4* expression have shorter overall survival when compared with patients with other expression groups. *Golim4* low expression was closely associated with advanced FIGO stage, higher histological grade, tumor size, and survival time. Semi-parametric Cox proportional hazards model showed that the low expression of Golim*4* and advanced FIGO stage were independent predictor of overall survival. In this study, the molecular mechanisms of *Golim4* were not studied in patients with EOC. Therefore, further studies are needed to prove the prognostic value of this gene.

## Data Availability

All data produced in the present work are contained in the manuscript

## Notes

### Competing Interest Statement

The authors have declared no competing interest.

### Funding Statement

This study did not receive any funding

### Author Declarations

Ethics committee of Tarbiat Modares university gave ethical approval for this work

